# Dual-Energy CT Angiography for Identifying High-Risk Carotid Plaques Associated With Ipsilateral Acute Ischemic Stroke: A Retrospective Case-Control Study

**DOI:** 10.1101/2025.09.08.25335142

**Authors:** Haoyue Shao, Yangyang Yin, Suping Chen, Hongyan Nie, Huan Liu, Lihui Dai, Yujie Zhao, Sikang Gao, Yanjie Zhao, Qiuxia Wang, Jing Zhang

## Abstract

**Background:** Early identification of high-risk carotid plaques is critical for acute ischemic stroke (AIS). Dual-energy CT angiography (DECTA) shows potential in quantifying high-risk plaque composition.

**Methods:** Patients with carotid plaques containing non-calcified components were retrospectively selected from 1,108 patients who underwent DECTA between January and November 2024. Maximum plaque thickness (MPT), CTA-based carotid Plaque-RADS classification, and DECTA parameters, including iodine concentration (IC), slope of the spectral curve (k), and effective atomic number (Z_eff_), were recorded. Differences were analyzed between the ipsilateral AIS (IAIS) plaques and the control plaques without ipsilateral symptom. Logistic regression analyses were performed to identify factors associated with IAIS. Least absolute shrinkage and selection operator (LASSO) regression was used to construct an optimal diagnostic model.

**Results:** Ninety-eight patients (mean age 62.0±11.1 years, 26.5% female) with 109 plaques were enrolled. Higher Z_eff_ (OR = 2.31, 95% CI: 1.09–5.19, p = 0.033) was identified as an independent factor associated with IAIS. Z_eff_ achieved higher area under the receiver operation characteristic curves (AUC) (95%CI) of 0.758 (0.670-0.847) than Plaque-RADS classification of 0.676 (0.577-0.775), although differences were not statistically significant (p > 0.05). The combined model further improved the AUC to 0.823 (95%CI: 0.744-0.901), significantly higher than Plaque-RADS (p = 0.002) and MPT (p < 0.001), while not statistically different from Z_eff_ (p > 0.05).

**Conclusions:** DECTA-derived parameters and the Plaque-RADS classification of carotid plaques were significantly correlated with IAIS. A higher Z_eff_ of non-calcified component was identified as an independent plaque feature associated with IAIS.

## Introduction

Carotid atherosclerosis is a well-established contributor to acute ischemic stroke (AIS) ^1,2^, with risk stratification traditionally based on luminal stenosis ^3–5^. However, over 50% of AIS cases and 55% of those with embolic stroke of undetermined source (ESUS) had non-stenotic plaques, highlighting the limited predictive value of stenosis alone ^6^. In decades, emerging studies suggested that plaque vulnerability is a more critical determinant of stroke risk, and it has traditionally been associated with its non-calcified components. Thus, identifying high-risk features in non-calcified components is crucial in facilitating earlier intervention and enhancing stroke prevention strategies ^7–9^.

Compositional and morphological plaque features, such as intraplaque hemorrhage (IPH), large lipid-rich necrotic cores (LRNC), and fibrous cap (FC) rupture, are strongly associated to plaque vulnerability and embolic potential ^7–9^. Recently, a structured imaging report system namely Carotid Plaque Reporting and Data System (Plaque-RADS) along with a CTA-based Plaque-RADS have emerged to generalize the plaque risk stratification process based on these plaque features ^10^. Although the clinical relevance of the Plaque-RADS classification has been preliminarily demonstrated, detailed subclassification requires high-resolution vessel wall imaging (HRVWI), and the semiquantitative classification results may be affected by subjective bias such as experience and energy ^11,12^. With the advances in CT technologies, dual-energy CT angiography (DECTA) providing quantitative parameters such as iodine concentration (IC), fat volume fraction (FVF), and effective atomic number (Z_eff_), have shown potential in recognizing LRNC, fibrous tissue, and IPH within the plaque ^13,14^. However, current research on DECTA-based plaque evaluation has been focused on isolated parameters and plaque component analysis ^15^, with limited efforts to validate their clinical value in identifying stroke-associated high-risk carotid plaques. Moreover, as CTA-based Plaque-RADS has recently been proposed, the comparison of Plaque-RADS and DECTA parameters in identifying high-risk plaques associated with AIS remains unexplored.

This study aimed to evaluate the value of DECTA–derived parameters and the Plaque-RADS classification in identifying high-risk plaque features associated with AIS in ipsilateral carotid plaques containing non-calcified components. We also attempted to develop a relatively comprehensive tool to support early identification of high-risk plaques and hoped to provide insights into developing personalized prevention strategies.

## Methods

### Patient Population

The study procedures were approved by the Institutional Review Board (IRB) of Tongji Hospital, Tongji Medical College, Huazhong University of Science and Technology before initiation. (TJ-IRB202503032). This retrospective study consecutively included 1,108 patients who underwent “one-stop CT imaging” for stroke (comprising non-contrast CT, head and neck DECTA, and brain CT perfusion), as well as head and neck DECTA between January and November 2024.

As shown in Figure 1, the inclusion criteria included: (1) patients with plaque in the extracranial segment of the carotid artery; (2) the plaque thickness >1mm; (3) AIS or no symptom ipsilateral to the plaque confirmed by CT or MR imaging ^16^.

**Figure 1.**
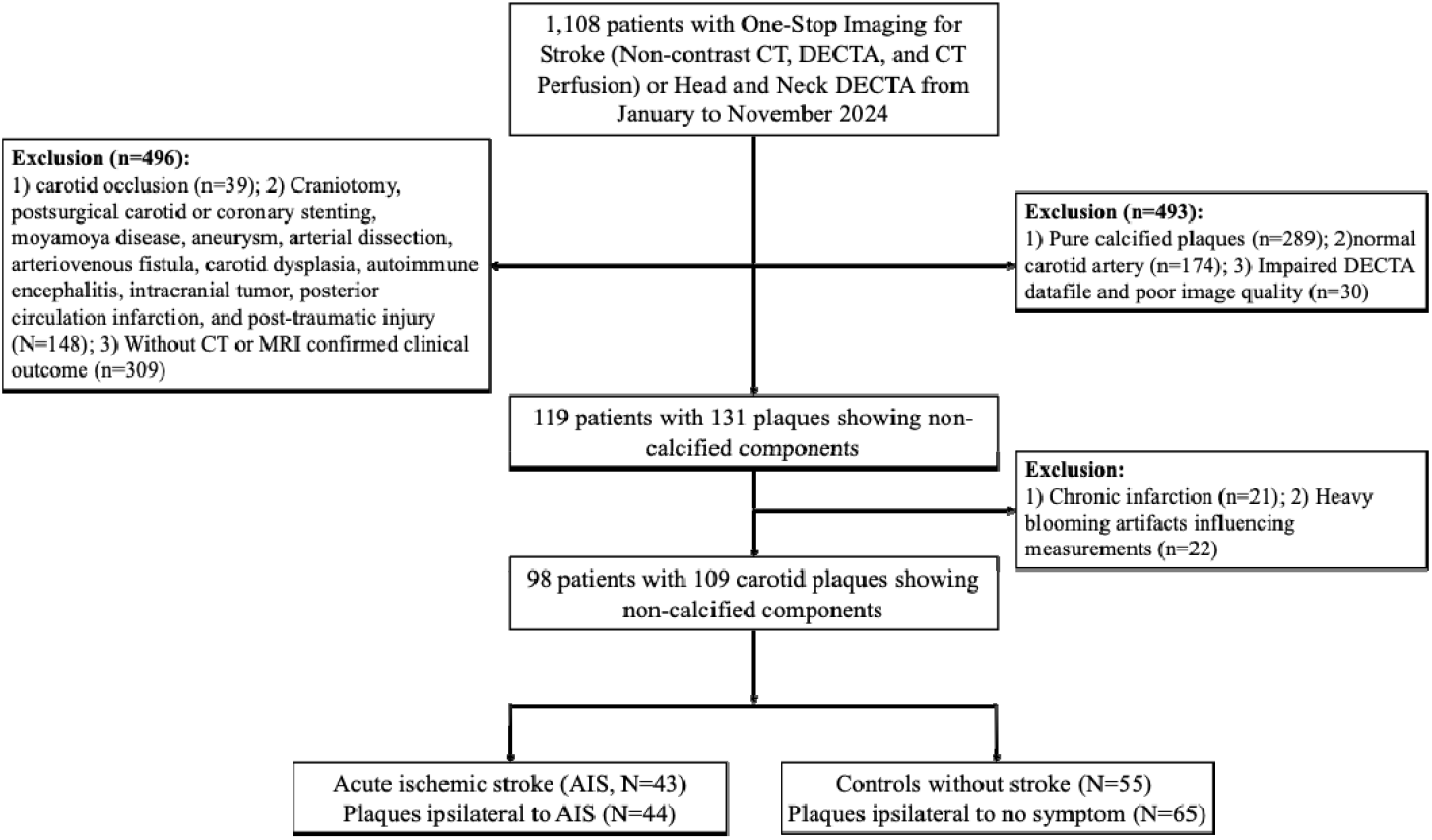
Flow diagram of patient selection. This diagram illustrates the inclusion and exclusion process for selecting patients and plaques for final analysis.

Exclusion criteria included: (1) occlusion of the carotid artery; (2) chronic infarction; (3) moyamoya disease, aneurysm, arterial dissection, arteriovenous fistula, carotid fibromuscular dysplasia, autoimmune encephalitis, intracranial tumor, and posterior circulation infarction; (4) post-surgical carotid or coronary stenting and post-traumatic injury; (5) pure calcified plaque; (6) plaque with heavy blooming artefacts influencing the measurement; (7) impaired DECTA data; (8) poor image quality.

### Clinical data

Comprehensive clinical data were obtained from the Hospital Information System (HIS) for each patient, including demographic information (age, gender, and BMI), relevant medical history (smoking status, alcohol consumption, hypertension, diabetes, and hyperlipidemia), and detailed biochemical indexes. These biochemical markers analyzed comprised triglycerides (TG), total cholesterol (TC), high-density lipoprotein cholesterol (HDL-C), low-density lipoprotein cholesterol (LDL-C), creatinine, and uric acid (UA).

### DECT angiography protocol

Head and neck DECTA was performed using a 256-row CT scanner (Revolution CT Power, GE HealthCare). Scan parameters: fast tube voltage switching between 80 kV and 140 kV, tube current: mean 449 mA, helical pitch: 0.992, scan range: aortic arch to the top of the skull. Reconstruction algorithm: adaptive statistical iterative reconstruction −Veo with a blending ratio of 50%, standard reconstruction kernel. Slice thickness: 0.625 mm. The contrast agent (370mgI/mL) with dosage based on the patient’s body mass index was injected through the right median cubital vein, and the injection time was fixed for 10 s. After the contrast injection, 20 ml of normal saline was injected at the same flow rate. The monitoring region of interest (ROI) was set at descending aorta and the scanning started at 2.5 seconds after the ROI attenuation reached 190 HU. The CT volume dose index was 8.73 mGy. Iodine-based maps, fat volume fraction images, effective atomic number images and 40-90keV virtual monochromatic images (VMIs) were generated for further evaluation and measurements.

### Carotid plaque image analysis

The included plaques were assessed through 100 kVp-like images. A neuroradiologist with 5 years of experience evaluated these plaques’ characteristics (mixed plaques or soft plaques) and measured stenosis degree (SD), maximum plaque thickness (MPT), and maximum plaque thickness of the non-calcified portion (MPT_NC_) according to the criteria of the North American Symptomatic Carotid Endarterectomy Trial (NASCET) ^17^, while blinded to the clinical outcome. Plaque grading was performed using the Carotid Plaque-RADS classification System ^10^. Plaque-RADS 2 is defined as low-risk plaque, 3a is defined as medium-risk plaque, 3c and 4 are defined as high-risk plaques.

For each plaque, three circular ROIs with areas of 0.5-1.5mm^2^ were delineated at the low-density area within the non-calcified components of the plaque and three adjacent slices, one superior and one inferior to the reference section (Figure 2). For each plaque, the non-calcified component was identified with CT attenuation <130 HU on 100-kVp like images ^18^. Within the non-calcified component, the lowest-attenuation area was manually selected for ROI placement, avoiding blooming artifacts. The HU window and location was adjusted to better display the area with the lowest attenuation relative to other parts in the selected plaque. The ROIs were then cloned to other DECT images to measure DECTA-derived parameters, including the CT values at 40 keV (HU_40keV_) and 90 keV (HU_90keV_), fat volume fraction (FVF), effective atomic number (Z_eff_), and iodine concentration (IC). Slope of the spectral attenuation curve (k) was calculated as following: k = (HU_40keV_ – HU_90keV_) /50. The mean values derived from these three ROIs were utilized for subsequent statistical analysis. For inter-consistency evaluation, thirty cases were randomly selected and another neuroradiologist with 10 years of experience performed Plaque-RADS classification and DECT measurements independently, blinded to the clinical outcome.

**Figure 2.**
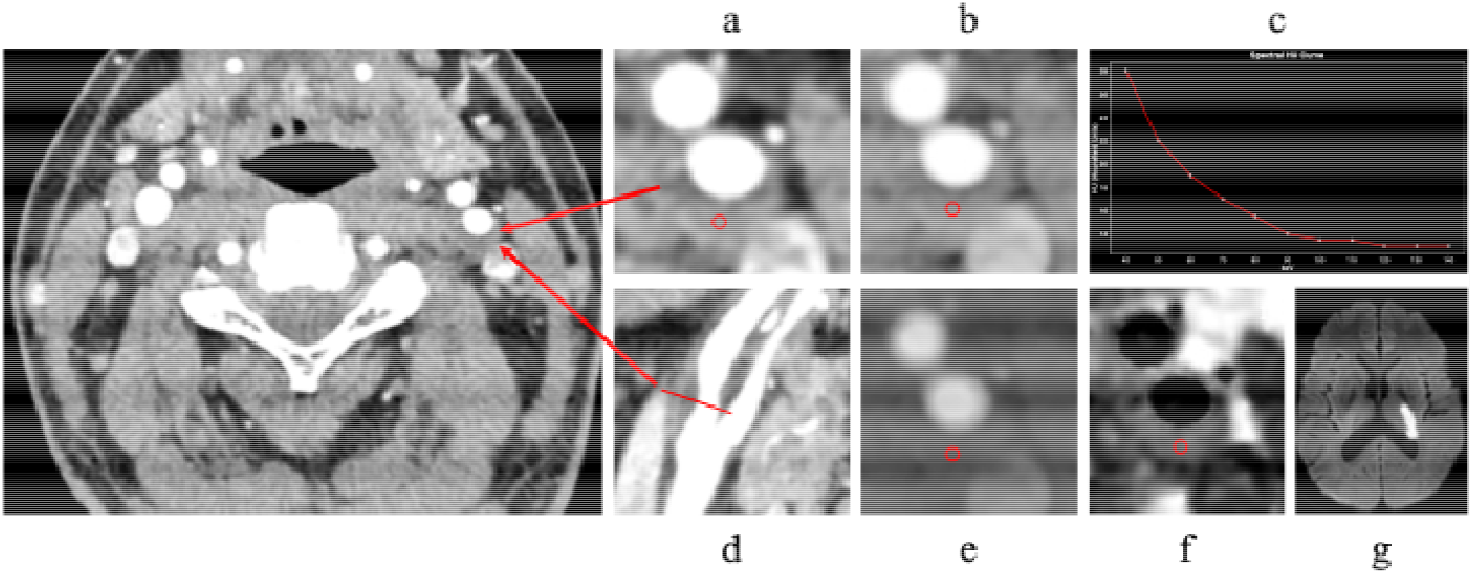
A middle-aged man was admitted to the hospital with slurred speech and right limb weakness for 7 hours. Representative DECTA image of a left carotid artery plaque classified as Plaque-RADS 3a. Diffusion-weighted images (DWI) show an ipsilateral acute ischemic stroke and the slope of the spectral curve shows a decreasing trend. a, Monoenergetic image of the axial position; b, Parametric image of effective atomic number (Z_eff_); c, The slope of the spectral Hounsfield unit curve (k); d, Image of Multi-Planar Reconstruction (MPR); e, Parametric image of iodine concentration (IC); f, Parametric image of fat volume fraction (FVF); g, Diffusion-weighted image. The region of interest (ROI) is manually delineated along the non-calcified component of the plaque for parameter analysis, including Z_eff_, k, IC, and FVF.

### Statistical analysis

Statistical analyses were performed using R (version 4.4.3; R Foundation for Statistical Computing, Vienna, Austria). Comparisons of continuous variables were conducted using either the Mann-Whitney U test or the independent samples t-test. All categorical variables were subject to the chi-square test or Fisher’s exact test. Univariate and multivariable logistic regression analyses were conducted to identify independent variables significantly associated with IAIS. Variables from the univariate logistic regression results (p□ <□ 0.1) were subsequently entered into a least absolute shrinkage and selection operator (LASSO) regression to construct a combined model. The performance of combined model and variables in diagnosing IAIS were assessed through receiver operating characteristic (ROC) curves. Delong’s test was used to compare the area under the ROC curves (AUC). A p-value < 0.05 was regarded as statistically significant. Interobserver agreement was evaluated using intraclass correlation coefficients (ICCs) for continuous variables (two-way random-effects model, absolute agreement) and Cohen’s kappa (κ) for categorical variables.

## Results

### Patient Characteristics

Of the 1108 screened patients, 98 patients (109 carotid plaques) were finally included. Forty-three AIS patients (five female, mean age 64.14 ± 9.66 years) with forty-four plaques ipsilateral to AIS (IAIS), fifty-five non-AIS individuals (twelve female, mean age 60.36 ± 11.97) with sixty-five control plaques without ipsilateral symptom. Detailed demographic, clinical, characteristics were summarized in Table 1.

**Table 1.** Demographic And Clinical Characteristics of Patients.

| Characteristics | Value |
| --- | --- |
| Patients (n) | 98 |
| Gender (Female, n, %) | 26 (26.5%) |
| Age (years, mean ± SD) | 62.0±11.1 |
| BMI (kg/m<sup>2</sup>, mean ± SD) | 24.1±3.7 |
| Smoking (n, %) | 24 (24.5%) |
| Drinking (n, %) | 18 (18.4%) |
| Hypertension (n, %) | 58 (59.2%) |
| Dyslipidemia (n, %) | 25 (25.5%) |
| Diabetes (n, %) | 10 (10.2%) |
| TC (abnormal, n, %) | 12 (15.2%) |
| TG (abnormal, n, %) | 17 (26.6%) |
| HDL-C (abnormal, n, %) | 34 (54.0%) |
| LDL-C (abnormal, n, %) | 12 (19.0%) |
| Creatinine (abnormal, n, %) | 30 (33.3%) |
| UA (abnormal, n, %) | 38 (46.3%) |

Values are presented as mean ± standard deviation or number (%).
Abbreviations: TC, total cholesterol; TG, triglycerides; HDL-C, high-density lipoprotein cholesterol; LDL-C, low-density lipoprotein cholesterol; UA, uric acid.

### Differences in RADS and DECTA Parameters Between IAIS and the Control plaques

The imaging characteristics between IAIS and control plaques were compared, as shown in Table 2. The IAIS plaques had higher-risk Plaque-RADS classification than those for controls (p < 0.001), while MPT, MPT_NC_, PL, and SD were not statistically different (all p > 0.05). As for DECTA-derived parameters, k, Z_eff_, and IC were significantly higher for IAIS plaques compared to those for controls (all p < 0.001), whereas FVF did not show statistically significant difference between the two groups (p = 0.096).

**Table 2.** Comparison of DECTA-derived parameters and Plaque-RADS features between plaques ipsilateral to acute ischemic stroke and ipsilateral to non-stroke.

| Variables | Ipsilateral to acute ischemic stroke<br>(n=44) | Ipsilateral to non-stroke (n=65) | *p* |
| --- | --- | --- | --- |
| MPT (mm) | 4.57 ± 2.04 | 3.81 ± 1.31 | 0.082 |
| MPT<sub>NC</sub> (mm) | 4.01 ± 2.15 | 3.40 ± 1.35 | 0.227 |
| PL (mm) | 24.09 ± 16.32 | 21.24 ± 17.51 | 0.105 |
| SD (%) | 0.24 ± 0.24 | 0.20 ± 0.15 | 0.826 |
| FVF (%) | 31.68 ± 11.79 | 29.32 ± 13.98 | 0.096 |
| IC (mg/cm<sup>3</sup>) | 11.52 ± 4.72 | 8.47 ± 3.79 | <0.001 |
| k | 1.02 ± 1.23 | 0.15 ± 0.99 | <0.001 |
| Z<sub>eff</sub> | 8.10 ± 0.45 | 7.69 ± 0.48 | <0.001 |
| Plaque characteristic |  |  | 0.399 |
| Mixed plaques | 32 (72.7%) | 41 (63.1%) |  |
| Soft plaques | 12 (27.3%) | 24 (36.9%) |  |
| RADS (n, %) |  |  | 0.001 |
| Low Risk | 9 (20.5%) | 18 (27.7%) |  |
| Medium Risk | 14 (31.8%) | 37 (56.9%) |  |
| High Risk | 21 (47.7%) | 10 (15.4%) |  |

Values are presented as mean ± standard deviation or number (%).
*P* values were calculated using the Student's t-test or Mann–Whitney U test for continuous variables and the $\chi^2$ test for categorical variables.
Abbreviations: MPT, maximum plaque thickness; MPT<sub>NC</sub>, maximum plaque thickness of the non-calcified portion; PL, plaque length; SD: vascular stenosis degree; FVF: Fat volume fraction in low-density areas of plaques; IC, iodine concentration; k, slope of spectral curve; Z<sub>eff</sub>, effective atomic number; Plaque-RADS, plaque reporting and data system. Low Risk, Plaque-RADS 2; Medium Risk, Plaque-RADS 3a; RADS High Risk, Plaque-RADS 3c and 4.

### Risk Factors Associated with IAIS

Higher value of MPT, k, Z_eff_, IC, and high-risk Plaque-RADS classification were significantly correlated with IAIS (all p < 0.05). In the multivariate logistic regression analysis, higher Z_eff_ was identified as a significant independent risk factor (2.31, 95% CI: 1.09-5.19, p = 0.033) (Figure 3).

**Figure 3.**
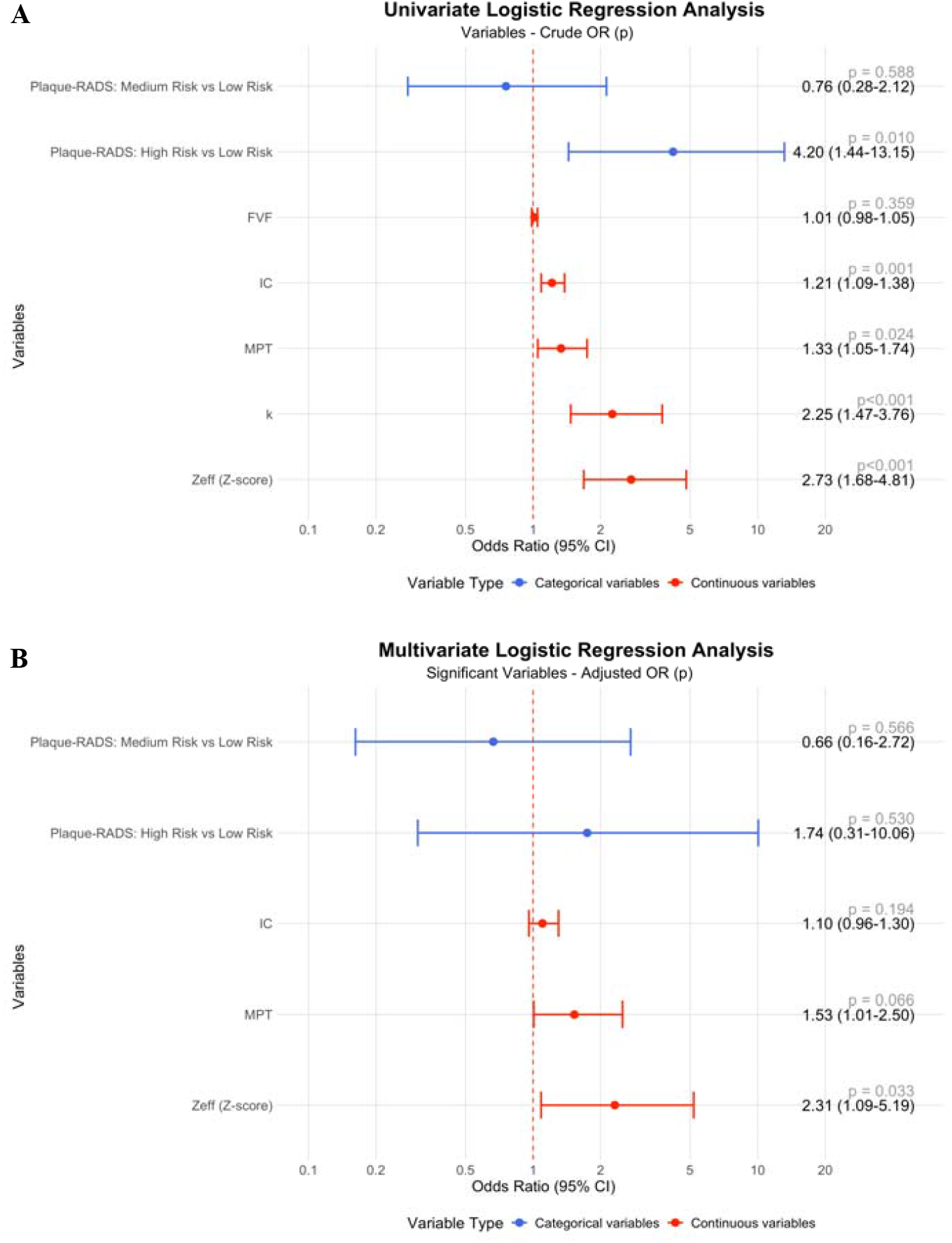
Forest plot illustrating the association of variables with outcome. A: Crude odds ratios (ORs) with 95% confidence intervals (CIs) are shown for univariable analyses. B: Adjusted ORs with 95% CIs for multivariable analyses. Variables included Plaque-RADS categories (Medium risk vs. Low risk; High risk vs. Low risk), FVF, IC, MPT, k, and Z_eff_ (Z-score). Odds ratios (ORs) and 95% confidence intervals (CIs) were calculated using logistic regression analysis. Variables with *p <* 0.10 in univariate analysis were included in the multivariate model. *P <* 0.05 was considered statistically significant. Abbreviations: OR, odds ratio; CI, confidence interval; Plaque-RADS, plaque reporting and data system score; Low Risk, Plaque-RADS 2; Medium Risk, Plaque-RADS 3a; High Risk, Plaque-RADS 3c and 4; MPT, maximum plaque thickness; FVF, fat volume fraction; IC, iodine concentration; k, slope of spectral curve; Z_eff_ (Z-score), standardized effective atomic number.

### Diagnostic performance of DECTA parameters and Plaque-RADS classification in identifying ipsilateral AIS-associated high-risk carotid plaques

In identifying IAIS-associated high-risk carotid plaques, the AUC was 0.676 (95% CI, 0.577-0.775) for the Plaque-RADS classification, 0.599 (0.485-0.713) for MPT, 0.749 (95% CI, 0.660-0.839) for IC, 0.768 (95% CI, 0.681-0.855) for k, and 0.758 (95% CI, 0.670-0.847) for standardized Z_eff_. Delong’s test indicated no significant difference in the AUC values between the Plaque-RADS classification and any DECTA parameters (Plaque-RADS vs. k, p = 0.154; Plaque-RADS vs. Z_eff_, p = 0.187; Plaque-RADS vs. IC, p = 0.272). The ROC analysis metrics are summarized in Table 3 and Figure 4.

**Table 3.** Summary of Diagnostic Performance Metrics.

| Variables | AUC (95%CI) | Sensitivity | Specificity | PPV | NPV | Accuracy | AIC | BIC |
| --- | --- | --- | --- | --- | --- | --- | --- | --- |
| Plaque-RADS classification | 0.676 (0.577-0.775) * | 47.73% | 84.62% | 67.74% | 70.51% | 69.72% | 139.30 | 147.38 |
| MPT | 0.599 (0.485-0.713) † | 45.45% | 81.54% | 62.50% | 68.83% | 66.97% | 145.43 | 150.82 |
| k | 0.768 (0.681-0.855) ‡ | 77.27% | 66.15% | 60.71% | 81.13% | 70.64% | 134.73 | 140.11 |
| IC | 0.749 (0.660-0.839) § | 93.18% | 50.77% | 56.16% | 91.67% | 67.89% | 137.31 | 142.69 |
| Z<sub>eff</sub> (Z-score) | 0.758 (0.670-0.847) \|\| | 90.91% | 53.85% | 57.14% | 89.74% | 68.81% | 132.10 | 137.48 |
| Combined model | 0.823 (0.744-0.901) | 77.27% | 75.38% | 68.00% | 83.05% | 76.15% | 122.78 | 138.93 |

Area under the curve (AUC) values with 95% confidence intervals (CI), along with sensitivity, specificity, positive predictive value (PPV), negative predictive value (NPV), overall accuracy, Akaike information criterion (AIC), and Bayesian Information Criterion (BIC) are shown for each model. The combined model included Plaque-RADS, MPT, k and Z<sub>eff</sub> (Z-score). \*, Plaque-RADS classification vs Combined model (p = 0.002); †, MPT vs Combined model (p < 0.001); ‡, k vs Combined model (p = 0.127); §, IC vs Combined model (p = 0.055); ||, Z<sub>eff</sub> (Z-score) vs Combined model (p = 0.074).
Abbreviations: Plaque-RADS, plaque reporting and data system score; MPT, maximum plaque thickness; IC, iodine concentration; k, slope of spectral curve; Z<sub>eff</sub> (Z-score), standardized effective atomic number.

**Figure 4.**
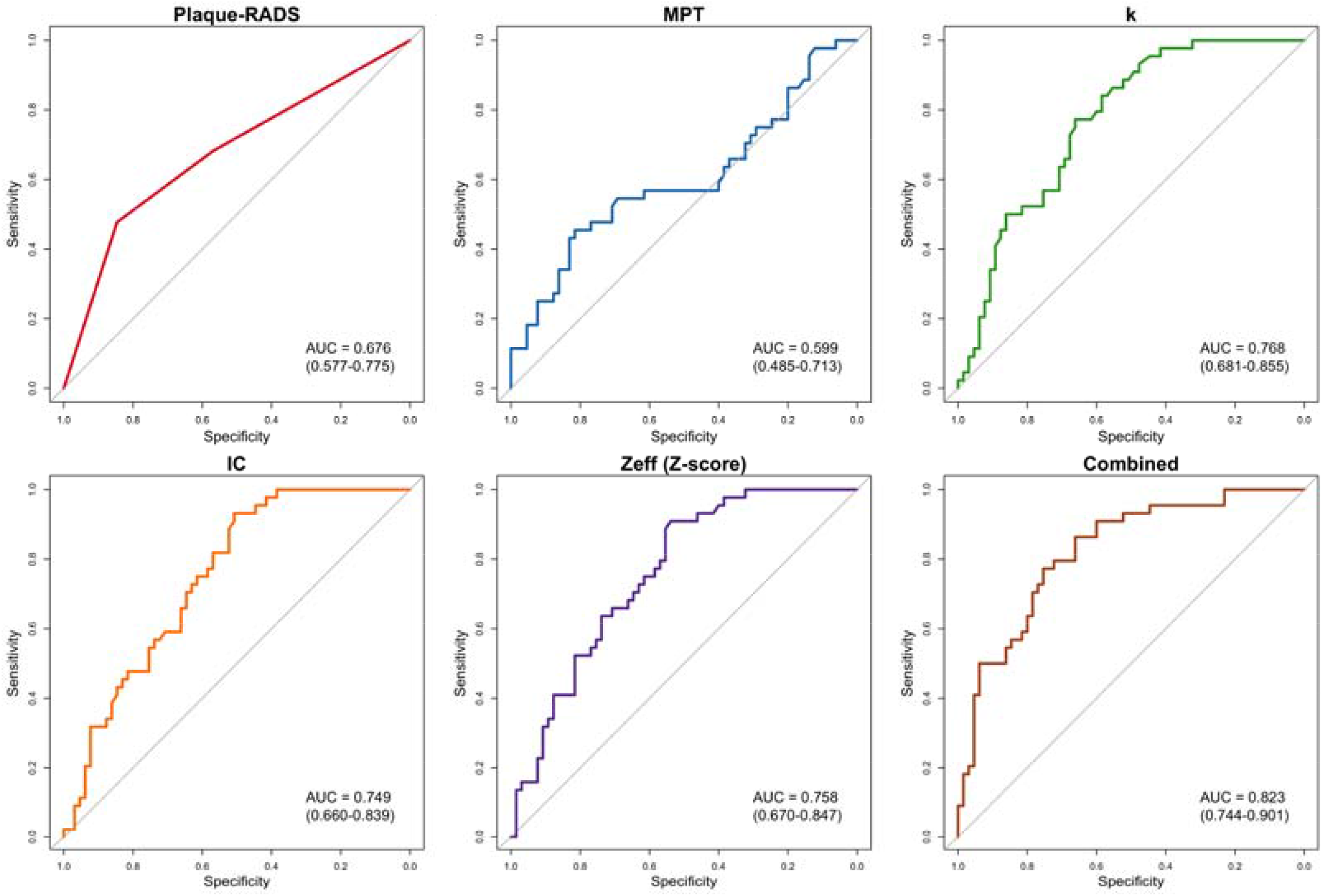
Receiver operating characteristic (ROC) curves were generated for five variables (Plaque-RADS, MPT, k, IC and Z_eff_) and a combined model integrating Plaque-RADS, MPT, k and Z_eff_. Abbreviations: Plaque-RADS, plaque reporting and data system score; MPT, maximum plaque thickness; k, slope of spectral curve; IC, iodine concentration; Z_eff_ (Z-score), standardized effective atomic number.

### Diagnostic performance of independent factors in identifying IAIS

The LASSO model incorporating Plaque-RADS, MPT, k and standardized Z_eff_ achieved an AUC of 0.823 (95% CI, 0.744-0.901), with Delong’s test p of 0.002 and p < 0.001 compared to Plaque-RADS classification and MPT, respectively. The model showed no statistically different AUC from DECTA parameters (all Delong’s test p > 0.05). The combined model showed 77.27% sensitivity, 75.38% specificity, 76.15% accuracy, as well as 68.00 % PPV and 83.05% NPV, at the optimal threshold. The model also demonstrated the optimal Akaike Information Criterion (AIC, 122.78) and Bayesian Information Criterion (BIC, 138.93) among all classifiers, indicating superior diagnostic performance and model fitting ability (Table 3 and Figure 4).

### Interobserver agreement of Plaque-RADS classifications and DECTA parameters

The weighted Cohen’s Kappa analysis revealed a Kappa value of 0.858 (p < 0.001) between the two radiologists for Plaque-RADS classification. Additionally, ICC values between the two radiologists were 0.928 for Z_eff_, 0.945 for k, 0.965 for IC, and 0.960 for FVF (p < 0.001).

## Discussion

Our study identified higher Z_eff_ of non-calcified component as an independent plaque factor associated with IAIS (OR = 2.31, 95% CI: 1.09-5.19, p = 0.033). Moreover, Z_eff_, k and IC derived from DECTA had higher AUC (0.758, 95% CI: 0.670-0.847; 0.768, 95% CI: 0.681-0.855; 0.749, 95% CI: 0.660-0.839, respectively] than CTA-based plaque-RADS classification [0.676 (0.577-0.775)] in identifying AIS-associated high-risk carotid plaques, showing no statistically significant difference though. The LASSO model integrating Plaque-RADS, MPT, IC and Z_eff_ achieved an AUC of 0.823 (95% CI, 0.744-0.901) in identifying IAIS associated high-risk carotid plaques.

Our study demonstrated higher Z_eff_, k, and IC values of the IAIS plaques than those of the asymptomatic controls, while Z_eff_ showed the strongest correlation with IAIS. The clinical significance of Z_eff_ lies in its ability to reflect tissue composition and provide valuable information for characterizing plaque vulnerability ^19–21^. Prior study demonstrated that the Z_eff_ showed a strong correlation with fibrous tissue, LRNC, and calcification, and a significant but weak correlation with IPH. Higher Z_eff_ value probably indicates increased microcalcification and contrast agent uptake, which correlate with features of plaque vulnerability, such as neovascularization, active inflammation, intraplaque hemorrhage, and fibrous cap rupture ^22^. However, Meng et al. reported that plaque Z_eff_ in the AIS group was significantly lower than that in the control group. This discrepancy might be attributed to the sample composition in their study, where calcified plaques were not excluded, and calcified plaques were more frequent in controls. Moreover, in their study, Z_eff_ measurements were performed on the whole plaque rather than non-calcified components. As a result, the measured Z_eff_ values could have been substantially influenced by the presence of calcified components, which typically elevated the Z_eff_ values. In addition, Zhang et al demonstrated that Z_eff_ in the symptomatic ischemic stroke group is also lower than that of the asymptomatic group. The possible reason for this difference may be that the symptomatic group in Zhang’s study included chronic infarction, which may exhibit different DECTA measurements ^23,24^. In addition, to avoid the influence of beam hardening artefacts of calcification components, the ROI area delineated in our study was smaller.

Additionally, the Plaque-RADS classification displayed by CTA offers a structured and repeatable framework for categorizing plaque morphology and we illustrated that the high-risk subtypes (3c and 4) showed correlations with IAIS, which was consistent with previous study ^25^. However, our results did not confirm the independent diagnostic value of the Plaque-RADS classification, which may be because the control group in our study was a non-AIS group, while the controls for Song’s study were plaques contralateral to AIS. In addition, fewer Plaque-RADS grade 2 and 4 patients were included in our study, and no Plaque-RADS grade 1 group. Wang’s research confirmed that the Plaque-RADS classification improved ischemic event risk evaluation in patients with mild and moderate stenosis ^11^. However, this study requires HRVWI to perform Plaque-RADS classification, and its application in patients with AIS is limited.

Our results found the independent value of Z_eff_ but not Plaque-RADS in identifying high-risk plaques. Our study demonstrated comparable or even higher diagnostic performance and better inter-reader agreement for DECTA parameters than Plaque-RADS classification. Nevertheless, there are also limitations to DECTA parameters for plaques with small low-density areas or significant calcified blooming artifacts, and Plaque-RADS classification remains applicable. Therefore, combining Plaque-RADS classification with DECTA parameters may provide broader clinical applicability. Our study further verified the improved diagnostic performance via the combination of Plaque-RADS, MPT, k and Z_eff_. This demonstrates that combining DECTA-derived plaque features and Plaque-RADS may provide a more comprehensive evaluation of plaque risk and guide personalized management strategies.

There are several research limitations worth considering in this study. First, a relatively small sample size and single-center design, which may restrict its generalizability. Second, the retrospective study is limited in preventing causal inferences, and a prospective cohort study is necessary to confirm the clinical predictive value. Third, the lack of external validation of the diagnostic models and CT scanner from a single manufacturer may affect the generalizability of our findings. Future research should focus on prospective multi-center validation, integrate artificial intelligence algorithms for automated plaque segmentation and DECTA parameter measurement, and conduct longitudinal follow-up to assess the prognostic value in predicting AIS.

Higher values of DECTA parameters such as Z_eff_, k, and IC, as well as higher-risk CTA-based RADS subtype of carotid plaques were correlated with IAIS, while Z_eff_ showed the highest correlation as an independent related factor. The model integrating Plaque-RADS, MPT, k and Z_eff_ may be a promising tool in AIS risk stratification, which needs further validation.

## Supporting information

checklist

## Data Availability

The data that support the results of this study are available from the corresponding author upon reasonable request. Individual participant data are not publicly available because of privacy and ethical restrictions.

## Acknowledgments

The authors sincerely thank the radiology technologists and nursing staff of the Department of Radiology, Tongji Hospital, for their assistance in patient data acquisition.

