## Supplementary material for "Dual-Energy CT Angiography for Identifying High-Risk Carotid Plaques Associated With Ipsilateral Acute Ischemic Stroke: A Retrospective Case-Control Study": checklist

STROBE Statement—checklist of items that should be included in reports of observational studies

|  | Item No. | Recommendation | Page  No. | Relevant text from manuscript |
| --- | --- | --- | --- | --- |
| **Title and abstract** | 1 | (*a*) Indicate the study’s design with a commonly used term in the title or the abstract | 1 | A Retrospective Case-Control Cohort Study |
|  |  | (*b*) Provide in the abstract an informative and balanced summary of what was done and what was found | 2 | A higher Z_eff_ of non-calcified component was identified as an independent plaque feature associated with IAIS |
| Introduction | | | |  |
| Background/rationale | 2 | Explain the scientific background and rationale for the investigation being reported | 3 | identifying high-risk features in non-calcified components is crucial in facilitating earlier intervention and enhancing stroke prevention strategies |
| Objectives | 3 | State specific objectives, including any prespecified hypotheses | 3 | develop a relatively comprehensive tool to support early identification of high-risk plaques and hoped to provide insights into developing personalized prevention strategies. |
| Methods | | | |  |
| Study design | 4 | Present key elements of study design early in the paper | 3 | DECTA and Plaque-RADS |
| Setting | 5 | Describe the setting, locations, and relevant dates, including periods of recruitment, exposure, follow-up, and data collection | 3 | 1,108 patients who underwent “one-stop CT imaging” for stroke (comprising non-contrast CT, head and neck DECTA, and brain CT perfusion), as well as head and neck DECTA |
| Participants | 6 | (*a*) *Cohort study*—Give the eligibility criteria, and the sources and methods of selection of participants. Describe methods of follow-up  *Case-control study*—Give the eligibility criteria, and the sources and methods of case ascertainment and control selection. Give the rationale for the choice of cases and controls  *Cross-sectional study*—Give the eligibility criteria, and the sources and methods of selection of participants | 3, 4 | AIS or no symptom ipsilateral to the plaque confirmed by CT or MR imaging |
|  |  | (*b*) *Cohort study*—For matched studies, give matching criteria and number of exposed and unexposed  *Case-control study*—For matched studies, give matching criteria and the number of controls per case | 3, 4 | 43 vs 55 |
| Variables | 7 | Clearly define all outcomes, exposures, predictors, potential confounders, and effect modifiers. Give diagnostic criteria, if applicable | 3 | AIS or no symptom ipsilateral to the plaque confirmed by CT or MR imaging |
| Data sources/ measurement | 8* | For each variable of interest, give sources of data and details of methods of assessment (measurement). Describe comparability of assessment methods if there is more than one group | 3, 4 | NASCET, CTA-based carotid Plaque-RADS classification |
| Bias | 9 | Describe any efforts to address potential sources of bias | 5 | intraclass correlation coefficients (ICCs) and Cohen’s kappa (κ) |
| Study size | 10 | Explain how the study size was arrived at | 3 | Patient Population |

Continued on next page

| Quantitative variables | 11 | Explain how quantitative variables were handled in the analyses. If applicable, describe which groupings were chosen and why | 3-5 | Statistical analysis  inclusion and exclusion criteria |
| --- | --- | --- | --- | --- |
| Statistical methods | 12 | (*a*) Describe all statistical methods, including those used to control for confounding | 5 | Statistical analysis |
|  |  | (*b*) Describe any methods used to examine subgroups and interactions | 5 | Statistical analysis |
|  |  | (*c*) Explain how missing data were addressed | 5 | Exclude missing data |
|  |  | (*d*) *Cohort study*—If applicable, explain how loss to follow-up was addressed  *Case-control study*—If applicable, explain how matching of cases and controls was addressed  *Cross-sectional study*—If applicable, describe analytical methods taking account of sampling strategy |  |  |
|  |  | (*e*) Describe any sensitivity analyses | 5 | Univariate and multivariable logistic regression analyses |
| Results | | | | |
| Participants | 13* | (a) Report numbers of individuals at each stage of study—eg numbers potentially eligible, examined for eligibility, confirmed eligible, included in the study, completing follow-up, and analysed | 3, 4 | inclusion and exclusion criteria |
|  |  | (b) Give reasons for non-participation at each stage | 3, 4 | inclusion and exclusion criteria |
|  |  | (c) Consider use of a flow diagram | 13 | Figure 1 |
| Descriptive data | 14* | (a) Give characteristics of study participants (eg demographic, clinical, social) and information on exposures and potential confounders | 4 | Clinical data |
|  |  | (b) Indicate number of participants with missing data for each variable of interest | 10 | Table 1 |
|  |  | (c) *Cohort study*—Summarise follow-up time (eg, average and total amount) |  |  |
| Outcome data | 15* | *Cohort study*—Report numbers of outcome events or summary measures over time |  |  |
|  |  | *Case-control study—*Report numbers in each exposure category, or summary measures of exposure | 11 | Table 2 |
|  |  | *Cross-sectional study—*Report numbers of outcome events or summary measures | 11 | Table 2 |
| Main results | 16 | (*a*) Give unadjusted estimates and, if applicable, confounder-adjusted estimates and their precision (eg, 95% confidence interval). Make clear which confounders were adjusted for and why they were included | 15 | Figure 3 |
|  |  | (*b*) Report category boundaries when continuous variables were categorized |  |  |
|  |  | (*c*) If relevant, consider translating estimates of relative risk into absolute risk for a meaningful time period |  |  |

Continued on next page

| Other analyses | 17 | Report other analyses done—eg analyses of subgroups and interactions, and sensitivity analyses | 5 | Risk Factors Associated with IAIS |
| --- | --- | --- | --- | --- |
| Discussion | | | | |
| Key results | 18 | Summarise key results with reference to study objectives | 6 | Higher Z_eff_ of non-calcified component as an independent plaque factor associated with IAIS |
| Limitations | 19 | Discuss limitations of the study, taking into account sources of potential bias or imprecision. Discuss both direction and magnitude of any potential bias | 7 | A relatively small sample size and single-center design |
| Interpretation | 20 | Give a cautious overall interpretation of results considering objectives, limitations, multiplicity of analyses, results from similar studies, and other relevant evidence | 6, 7 | This discrepancy might be attributed to the sample composition in their study, where calcified plaques were not excluded, and calcified plaques were more frequent in controls. Moreover, in their study, Z_eff_ measurements were performed on the whole plaque rather than non-calcified components.  In addition, to avoid the influence of beam hardening artefacts of calcification components, the ROI area delineated in our study was smaller. |
| Generalisability | 21 | Discuss the generalisability (external validity) of the study results | 7 | Future research should focus on prospective multi-center validation, integrate artificial intelligence algorithms for automated plaque segmentation and DECTA parameter measurement, and conduct longitudinal follow-up to assess the prognostic value in predicting AIS |
| Other information | |  | | |
| Funding | 22 | Give the source of funding and the role of the funders for the present study and, if applicable, for the original study on which the present article is based |  |  |

*Give information separately for cases and controls in case-control studies and, if applicable, for exposed and unexposed groups in cohort and cross-sectional studies.

**Note:** An Explanation and Elaboration article discusses each checklist item and gives methodological background and published examples of transparent reporting. The STROBE checklist is best used in conjunction with this article (freely available on the Web sites of PLoS Medicine at http://www.plosmedicine.org/, Annals of Internal Medicine at http://www.annals.org/, and Epidemiology at http://www.epidem.com/). Information on the STROBE Initiative is available at www.strobe-statement.org.
